# A phylogeny-aware GWAS framework to correct for heritable pathogen effects on infectious disease traits

**DOI:** 10.1101/2021.11.22.21266687

**Authors:** Sarah Nadeau, Christian W. Thorball, Roger Kouyos, Huldrych F. Günthard, Jürg Böni, Sabine Yerly, Matthieu Perreau, Thomas Klimkait, Andri Rauch, Hans H. Hirsch, Matthias Cavassini, Pietro Vernazza, Enos Bernasconi, Jacques Fellay, Venelin Mitov, Tanja Stadler, the Swiss HIV Cohort Study (SHCS)

**Affiliations:** Department of Biosystems Science and Engineering, ETH Zurich, Basel, Switzerland; Swiss Institute of Bioinformatics, Lausanne, Switzerland; Precision Medicine Unit, Lausanne University Hospital and University of Lausanne, Lausanne, Switzerland; Institute of Medical Virology, University of Zurich, Zurich, Switzerland; Division of Infectious Diseases and Hospital Epidemiology, University Hospital Zurich, University of Zurich, Zurich, Switzerland; Division of Infectious Diseases, Laboratory of Virology, Geneva University Hospital, Geneva, Switzerland; Division of Immunology and Allergy, University Hospital Lausanne, Lausanne, Switzerland; Department of Biomedicine, University of Basel, Basel, Switzerland; Department of Infectious Diseases, Bern University Hospital and University of Bern, Bern, Switzerland; Division of Clinical Virology, University Hospital Basel, Basel, Switzerland; Division of Infectious Diseases and Hospital Epidemiology, University Hospital Basel, Basel, Switzerland; Division of Infectious Diseases, University Hospital Lausanne, Lausanne, Switzerland; Division of Infectious Diseases, Cantonal Hospital St. Gallen, St. Gallen, Switzerland; Division of Infectious Diseases, Regional Hospital Lugano, Lugano, Switzerland; Global Health Institute, School of Life Sciences, École Polytechnique Fédérale de Lausanne, Lausanne, Switzerland

**Author notes:** Co-last authors (Venelin Mitov, Tanja Stadler).

## Abstract

Infectious diseases are a unique challenge for genome-wide association studies (GWAS) because pathogen, host, and environmental factors can all affect disease traits. Previous GWAS have successfully identified several human genetic variants associated with HIV-1 set point viral load (spVL), among other important infectious disease traits. However, these GWAS do not account for potentially confounding or extraneous pathogen effects that are heritable from donor to recipient in transmission chains. We propose a new method to consider the full genome of each patient’s infecting pathogen strain, remove strain-specific effects on a trait based on the pathogen phylogeny, and thus better estimate the effect of human genetic variants on infectious disease traits. In simulations, we show our method can increase GWAS power to detect truly associated host variants when pathogen effects are highly heritable, with strong phylogenetic correlations. When we apply our method to HIV-1 subtype B data from the Swiss HIV Cohort Study, we recover slightly weaker but qualitatively similar signals of association between spVL and human genetic variants in the *CCR5* and major histocompatibility complex (MHC) gene regions compared to standard GWAS. Our simulation study confirms that based on the estimated heritability and selection parameters for HIV-1 subtype B spVL, standard GWAS are robust to pathogen effects. Our framework may improve GWAS for other diseases if pathogen effects are even more phylogenetically correlated amongst individuals in a cohort.

## Introduction

A key goal of genome-wide association studies (GWAS) is to understand the genetic basis of phenotypic variation among individuals. In a typical GWAS, millions of genetic variants from across the human genome are screened for statistical association with a trait of interest. Ideally, this procedure identifies variants that are located in, or are in linkage disequilibrium with, alleles that directly affect the trait. If GWAS finds a variant strongly associated with a disease trait, the gene product may be a good drug target (Okada *et al*., 2014). Even if no single variant has a strong association, many small associations can be aggregated into a polygenic risk score to identify high-risk individuals (Dudbridge, 2013).

For HIV, GWAS have used a trait called set point viral load (spVL) to identify human variants associated with the severity of disease course. spVL is generally defined to be the average concentration of viral RNA copies in host plasma during the asymptomatic phase of infection in the absence of treatment (see e.g. Alizon *et al*. (2010)). In untreated individuals, spVL is predictive of the duration of asymptomatic infection (Mellors *et al*., 1996) and infectiousness (Quinn *et al*., 2000). If viral load can be reduced to undetectable levels, an individual is effectively uninfectious and the risk of disease progression is massively reduced (Panel on Antiretroviral Guidelines for Adults and Adolescents, 2019). Notably, spVL varies by orders of magnitude between individuals (Mellors *et al*., 1996). Thus, spVL measurements point to a wide range in natural HIV control amongst individuals.

To understand HIV pathogenicity, it is important to understand to what extent spVL is determined by host genetic factors (Bartha *et al*., 2013; Dalmasso *et al*., 2008; Fellay *et al*., 2007, 2009; McLaren *et al*., 2012; Pelak *et al*., 2010; Pereyra *et al*., 2010; van Manen *et al*., 2011). Heritability is a key measure of how genetically-determined a trait is. Here we distinguish between two different heritability measures that are used in different contexts in the study of infectious diseases. Broad-sense heritability *H*^2^ measures the fraction of total trait variance that is heritable, i.e. due to inherited differences. In the infectious disease case, broad-sense heritability from pathogen factors, which are inherited by recipients from their infection partners, is typically measured. On the other hand, narrow-sense heritability *h*^2^ measures the fraction of total trait variance due specifically to additive genetic effects, i.e. the sum of independent effects from all genetic variants. GWAS for infectious disease traits typically measure the narrow-sense heritability of a trait based on human genetic variants.

Several GWAS have been done to measure the narrow-sense heritability of spVL and identify associated host genetic variants (Bartha *et al*., 2013; Dalmasso *et al*., 2008; Fellay *et al*., 2007, 2009; McLaren *et al*., 2012; Pelak *et al*., 2010; Pereyra *et al*., 2010; van Manen *et al*., 2011). The largest study to-date by McLaren *et al*. (2015) estimated the narrow-sense heritability of spVL from human genetic variants to be approximately 25%. All but 5% of this was attributed to two regions in the human genome, the major histocompatibility complex (MHC) and C-C motif chemokine receptor 5 (*CCR5*). Both associations are biologically relevant: the MHC encodes proteins that present viral epitopes at the cell surface and *CCR5* encodes a co-receptor for HIV-1 cell entry. In other words, MHC proteins match bits of the virus like puzzle pieces and display these to signal that a cell is infected. CCR5 proteins help the virus infect target cells.

In addition to these human genetic factors, it is well-recognized that viral genetic factors affect spVL. As mentioned, heritability from the viral side is typically measured using broad-sense heritability. Estimates differ depending on the methods employed and the cohort studied (see Mitov and Stadler (2018) for a discussion of this uncertainty). Estimates using phylogenetic methods on large UK and Swiss cohorts by Mitov and Stadler (2018) and Bertels *et al*. (2018) measured the broad-sense heritability of spVL from the virus to be 21% - 29%. However, variation in the MHC is known to exert strong selective pressure on the virus (Kløverpris *et al*., 2016; Nguyen *et al*., 2021). If the virus can change its “puzzle piece” shape to escape MHC-presentation, infected cells can go undetected. This means that MHC variants affect spVL largely via selection on the virus (Bartha *et al*., 2017). In summary, human genetic factors play a role in determining spVL, but these effects may be due to interaction with specific viral genetic variants.

Most of the GWAS for human genetic determinants of spVL conducted so far (Dalmasso *et al*., 2008; Fellay *et al*., 2007, 2009; McLaren *et al*., 2012, 2015; Pelak *et al*., 2010; Pereyra *et al*., 2010; van Manen *et al*., 2011) do not explicitly consider any viral effect on spVL. In these GWAS, viral genetic effects are lumped in with residual variance due to other, non-genetic factors. This has several potential negative consequences. (i) Viral effects may be confounding or extraneous variables that bias estimates of host genetic effects. (ii) Variability due to viral effects would make it more challenging to identify human variants of small effect. Finally, (iii) spVL values from a cohort are not truly independent samples, given that patients closer in the transmission chain have more similar strains and therefore more similar spVL values.

Issue (iii) is closely related to a well-known problem in standard GWAS. Shared (human) ancestry, especially between close relatives, can also give rise to spurious genetic correlations with a trait. Corrections for these correlations are well-developed and widely accepted (Astle and Balding, 2009; Price *et al*., 2006). More recently, Power *et al*. (2017) emphasized the need to do similar corrections for shared pathogen ancestry in microbial GWAS. Two state-of-the-art methods exist for this (Collins and Didelot, 2018; Earle *et al*., 2016). However, these approaches are only suitable to quantify effects from pathogen genetic variants on a trait. In contrast, we want to estimate effects of human genetic variants on a trait, while accounting for pathogen effects. Naret *et al*. (2018) developed a relevant method for this in the context of a genome-to-genome GWAS framework. The authors suggest adding principle components derived from the pathogen phylogeny as covariates to the linear regression models for association testing. This should correct for trait correlations due to shared pathogen ancestry. However, the top principle components capture only some of the information from the full pathogen phylogeny. Furthermore, we would like to simultaneously address issues (i) and (ii).

In this work, we draw from the field of phylogenetic comparative methods to develop a new GWAS framework that estimates and removes trait variability due to the pathogen using information from the full pathogen phylogeny. Our approach should help identify human genetic variants that affect disease traits and more accurately estimate their effects.

In the following we describe a statistical model for the spVL trait, derive a maximum likelihood estimate for the viral part of spVL under this model, and describe a new infectious disease GWAS framework using this information. In simulations, we show that this framework can improve GWAS power to detect host genetic variants that affect disease traits. Finally, we apply our framework to human and viral genome data from the Swiss HIV Cohort Study (SHCS) and show that associations with spVL are robust to a correction for viral effects. Although we developed our framework in the context of HIV-1 spVL, this approach can readily be applied to other heritable infectious disease traits.

## New Approaches

### A statistical model for spVL

Variation in spVL comes from several sources: direct host genetic effects, pathogen effects, interaction effects between the host and the pathogen, and other environmental effects. Of these, only pathogen effects are heritable from one transmission partner to another (Leventhal and Bonhoeffer, 2016). To characterize these effects, we use a phylogenetic mixed model (PMM) (Housworth *et al*., 2004). PMMs assume continuous traits like spVL are the sum of independent heritable and non-heritable parts. In our case, pathogen effects comprise the heritable part and all other effects comprise the non-heritable part. The heritable part is modeled by a random process occurring in continuous time along the branches of the pathogen phylogeny, as in Figure 1A. The non-heritable part is modeled as Gaussian noise added to sampled individuals at the tips of the phylogeny.

**Figure 1:**
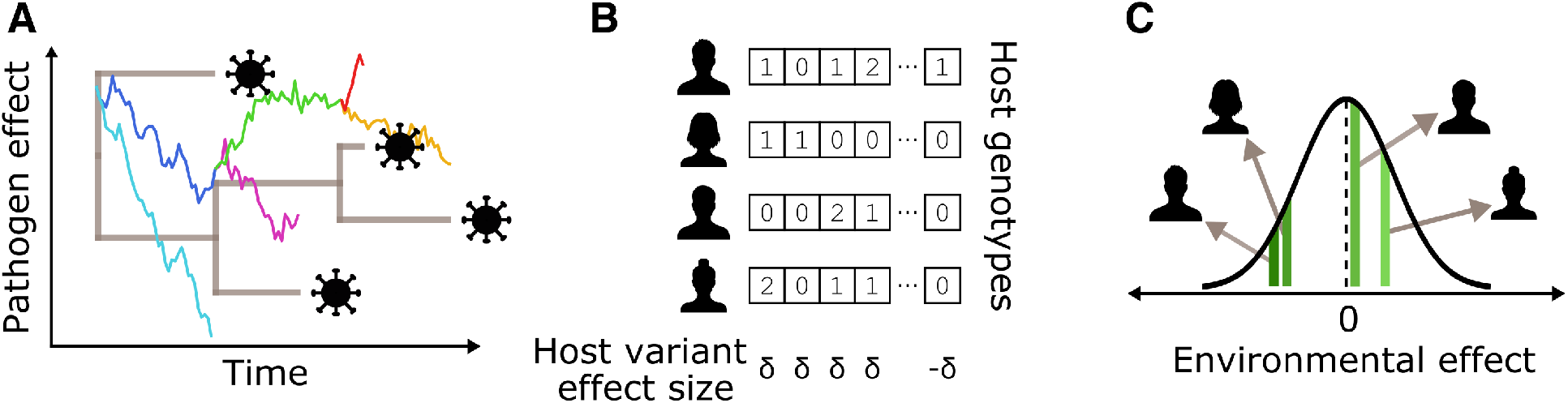
A high-level schematic of our POUMM-based simulation framework. (A) shows how pathogen genetic effects on spVL evolve along the pathogen phylogeny according to an OrnsteinUhlenbeck process. (B) shows how host genetic effects are the sum of independent effects from several causal variants. Each variant can be present in 0, 1, or 2 copies. Half the variants have a positive effect of size *δ* and half have a negative effect of size *δ*. (C) shows how environmental effects are independently drawn from a Gaussian distribution centered at 0. These three effects sum to the trait value for each simulated individual.

So far, PMMs with two types of random processes have been used to model spVL evolution. The Brownian Motion (BM) process assumes unbounded trait values, i.e. spVL can attain any value. The Ornstein-Uhlenbeck (OU) process assumes trait values fluctuate around an optimal value, i.e. extreme spVL values are unlikely. Mitov and Stadler (2018) and Bertels *et al*. (2018) previously showed the OU process has higher statistical support for spVL. This makes sense given that spVL is likely under stabilizing selection to maximize viral transmission potential (Fraser *et al*., 2014). Therefore, we assume the OU process. The full model is called the phylogenetic Ornstein-Uhlenbeck mixed model (POUMM) and is described in detail by Mitov and Stadler (2018). Here, we review the main points in the spVL context.

Under the POUMM, the spVL trait *z* is the sum of viral effects *g*_*v*_, host genetic effects *g*_*h*_, and other environmental or interaction effects *ϵ*. We can group the non-heritable effects *g*_*h*_ and *ϵ* into a broader category of “environmental” effects *e*:

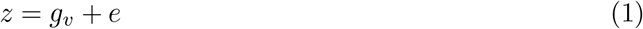

*g*_*v*_ is a viral trait that evolves along the phylogeny according to an OU process. The OU process is defined by a stochastic differential equation with two terms. The first term represents a deterministic pull towards an optimal trait value and the second term represents stochastic fluctuations modelled by Brownian motion (Butler and King, 2004):

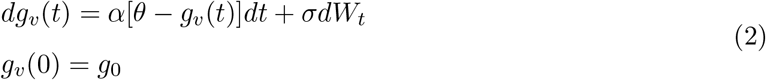

Here the parameter *α* represents selection strength towards an evolutionarily optimal value represented by parameter *θ*. The parameter *σ* measures the intensity of stochastic fluctuations in the evolutionary process. Finally, *dW*_*t*_ is the Wiener process underlying Brownian motion. The OU process is a Gaussian process, meaning that *g*_*v*_(*t*) is a Gaussian random variable. Assuming *g*_*v*_(*t*) starts at initial value *g*_*0*_ at time *t* = 0 at the root of the phylogeny, we can write the expectation for *g*_*v*_(*t*) at time *t:*

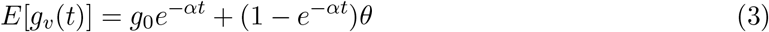

and the variance in *g*_*v*_(*t*) if we were to repeat the random evolutionary process many times (Butler and King, 2004):

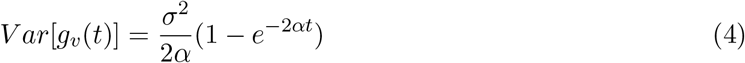

*g*_*v*_ evolves independently in descendent lineages after a divergence event in the phylogeny. The covariance between *g*_*v*_(*t*) in a lineage *i* at time *t*_*i*_ and another lineage *j* at time 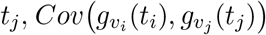, increases with the amount of time between *t*_0_ and the divergence of the two lineages, *t*_0(*ij*)_, and decreases with the total amount of time the lineages evolve independently, *d*_*ij*_ (Butler and King, 2004):

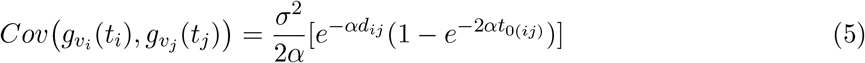

Next, we remember that *e* is the non-heritable, environmental part of spVL. *e* is modeled as a Gaussian random variable that is time- and phylogeny-independent. The expectation of *e* is 0, meaning environmental effects are equally likely to raise or lower spVL from the virus-determined level. The parameter 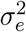 measures the between-host variance of the environmental effect.

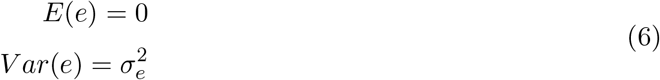

Finally, broad-sense trait heritability can be calculated as the fraction of total trait variance that is heritable:

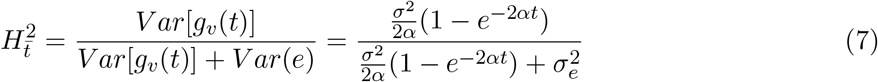

### Teasing apart pathogen and non-pathogen effects on spVL

Given the assumptions of the POUMM, we can estimate a heritable pathogen effect on spVL and a non-heritable, host and environmental effect on spVL. Here, we derive a maximum-likelihood estimate for these values for individuals in a cohort, given measured spVL values and a pathogen phylogeny linking the infecting strains.

Let *g*_*v*_(*t*) be a vector of *g*_*v*_ values, one for each individual in the cohort. *t* are the sampling times of each individual relative to the root of the phylogeny. To simplify notation, we omit the *t* from here on. *g*_*v*_ is a realization of a Gaussian random vector *G*_*v*_ ∼ 𝒩 (*μ*_*OU*_, Σ_*OU*_). The expectation *μ*_OU_ is defined by equation 3, the diagonal elements of the covariance matrix Σ_*OU*_ are defined by equation 4, and the off-diagonal elements of Σ_*OU*_ by equation 5.

Similarly, let *e* be a vector of the environmental part of spVL for each individual. *e* is a realization of a Gaussian random vector *E* ∼𝒩(0, Σ_*E*_), where Σ_*E*_ is a diagonal matrix with diagonal elements equal to 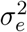.

Considering that *G*_*v*_ and *E* are independent random vectors and that their realizations *g*_*v*_ and *e* must sum together to equal the observed spVL values *z*, we can write the following proportionality for the joint probability density of *g*_*v*_ and *e*:

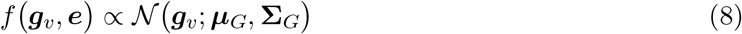

where the expected value of *g*_*v*_ and the covariance matrix **Σ**_*G*_ are defined as:

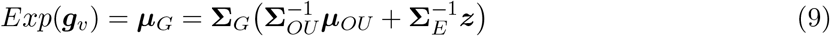

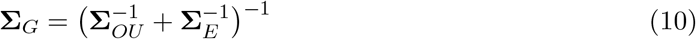

*Proof*.

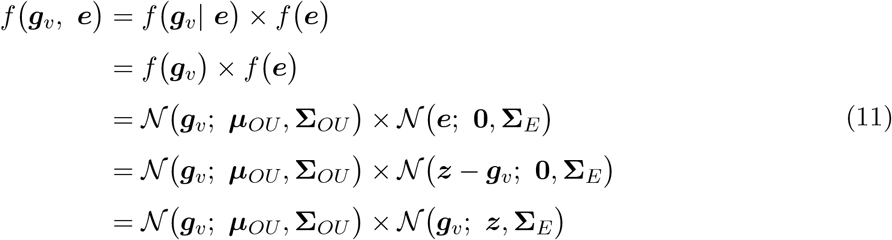

Equations 9 and 10 follow from eq. 11 and eq. 371, p. 42, section 8.1.8 “Product of Gaussian densities” in Petersen and Pedersen (2012).□

Importantly, equation 9 is the maximum likelihood estimate for *g*_*v*_, the viral effect on spVL, taking into account all available information - measured spVL, the pathogen phylogeny, and inferred POUMM parameters. This estimator is an inverse-variance weighted average of measured spVL and information from the POUMM evolutionary model (*μ*_OU_). In other words, *g*_*v*_ will be closer to measured spVL if spVL is not very heritable. If spVL is highly heritable, *g*_*v*_ will be closer to the expected value under the POUMM, i.e. take more information from the phylogenetic relationships between infecting strains.

Given the estimator we just derived for *g*_*v*_, we can now estimate *e*, the spVL value without pathogen effects:

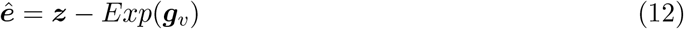

We will use this value to try to improve upon standard GWAS methods in infectious disease.

### A POUMM-based GWAS framework for infectious disease

We propose to improve standard GWAS for infectious diseases by estimating and removing trait variability due to pathogen effects. Our new framework is as follows:

1. Sample host genotypes, trait values, and pathogen genome sequence data from a cohort.
2. Construct a pathogen phylogeny using the pathogen genome sequences.
3. Estimate the parameters of the POUMM based on the trait values and the pathogen phylogeny. This can be done with e.g. the R package POUMM (Mitov and Stadler, 2017).
4. Generate maximum-likelihood estimates for the pathogen and corresponding non-pathogen effects on the trait using equations 9 and 12.
5. Perform GWAS with only the non-pathogen effects on the trait as the response variable.

## Results

### Simulation study

To test the theoretical best-case performance of our method, we simulated data under the POUMM and applied our framework to the simulated data. Figure 1 shows a high-level schematic of our simulation framework and Table 2 gives the value or expression for each parameter. For a description of the full simulation scheme, see Figure S1. In a nutshell, we simulated independent, additive host genetic effects, independent environmental effects, and heritable pathogen genetic effects under different scenarios of trait heritability and selection strength. To maintain the same heritability while varying selection strength, we counter-balanced by varying the intensity of stochastic evolutionary fluctuations accordingly. We fixed other variables to plausible values based on the spVL literature.

### Estimator accuracy

First, we evaluated how well our method estimated the additive host genetic effects from the simulated data. Additive host genetic effects represent an ideal (albeit unattainable) baseline for infectious disease GWAS. Figure 2A shows that our method incorporating phylogenetic information can more accurately estimate these value compared to the trait value. To ensure a fair comparison, we scaled trait values to have the same mean, zero, as host genetic effects so as not to bias the root mean squared error (RMSE) by a constant factor. In the supplemental material, we show why the scaled trait value is expected to have an RMSE of approximately 0.74 under our simulation scheme. By incorporating phylogenetic information, we can improve upon this error in scenarios where the trait is highly heritable, under low selection pressure, and with relatively moderate stochastic fluctuations compared to outbreak duration. This is because high heritability means the trait value is highly pathogen-dependent. Then, when the trait is under weak selection, the pathogen effects can drift far from the long-term optimum and stochastic fluctuations are low to maintain the same heritability. Thus, our method performs well when an infectious disease trait is highly heritable and trait values are strongly correlated amongst close transmission partners.

**Figure 2:**
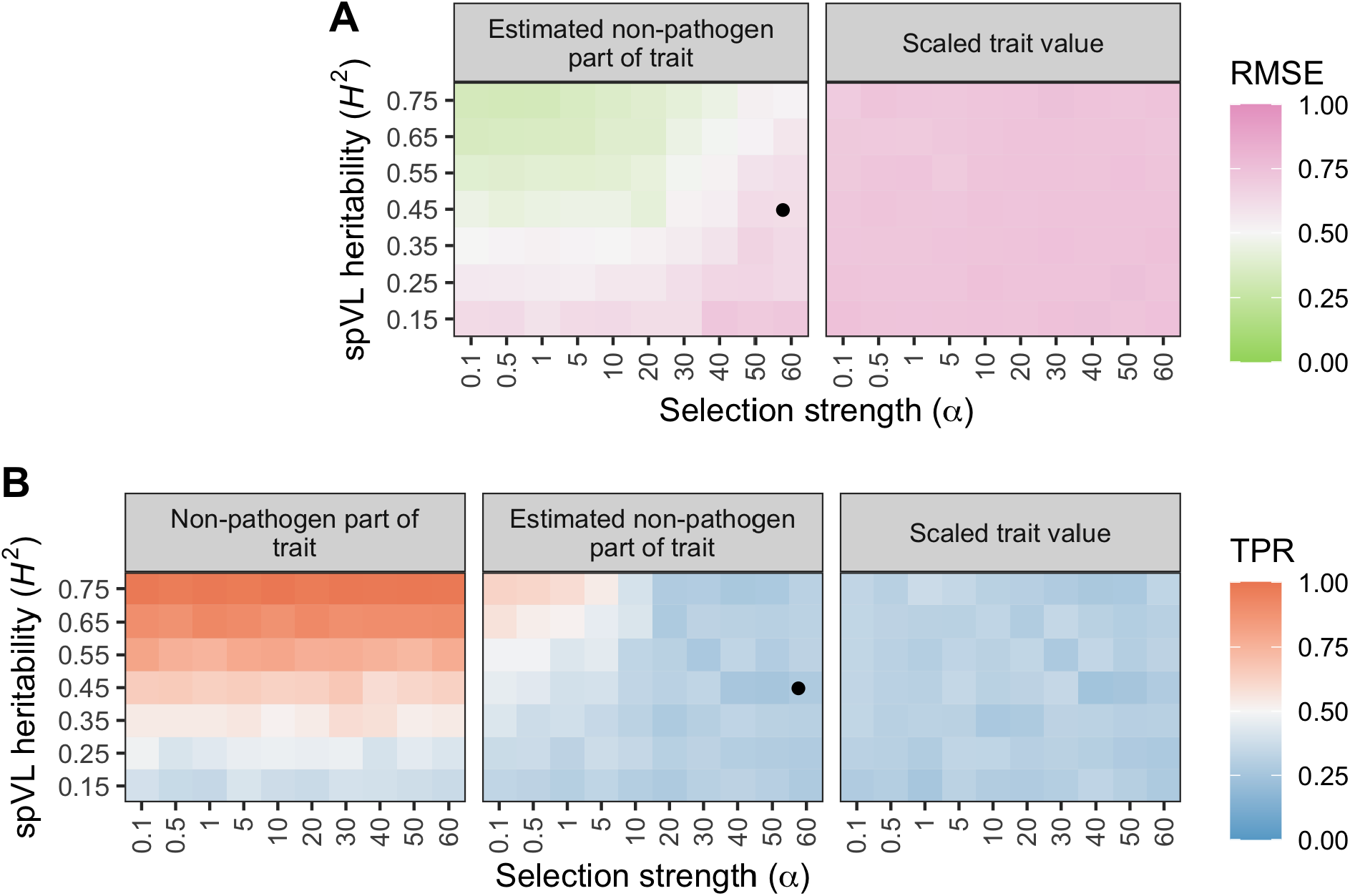
Results from the simulation study. We simulated host, pathogen, and environmental effects on a trait under the POUMM with different heritability (y-axis) and selection strength (xaxis) parameters. For each simulated dataset, we applied our method to estimate the non-pathogen effects and performed GWAS with these values. (A) shows that our method (left) can generate more accurate estimates of additive host genetic effects than the trait value, scaled by its mean (right). (B) shows how GWAS power can improve given the true, simulated non-pathogen effect on spVL (left) and using our estimate for this value (middle) compared to using the scaled trait value (right). Each tile’s color corresponds to the average value across 20 simulated datasets of 500 samples. The black point represents our estimates for the heritability and selection strength of spVL based on Swiss HIV Cohort Study data. RMSE = Root mean square error, TPR = True positive rate.

### Theoretical GWAS improvement

Next, we characterized the evolutionary scenarios under which our framework can actually improve GWAS power. We used the true positive rate (TPR) to evaluate the fraction of simulated causal host genetic variants we could recover as being significantly associated with the trait. We performed three different GWAS for each simulated dataset: the first represents an ideal in which we can exactly know and remove pathogen effects from trait values, the second is using our method to estimate this value and remove it, and the third represents a standard GWAS using the scaled trait value. Figure 2B shows that our framework can improve the TPR in simulated scenarios where selection strength < 10 time^−1^ and heritability > 45%. If we were able to perfectly estimate and remove pathogen effects from a trait, the TPR would increase across all values of selection strength so long as the trait is more than marginally heritable. We estimate approximately 25% to be the heritability threshold above which GWAS power is negatively impacted by pathogen effects. In summary, we show it is theoretically possible to improve GWAS power for heritable infectious disease traits by estimating and removing pathogen effects using information from the pathogen phylogeny.

### GWAS on the Swiss HIV Cohort

Finally, we applied our framework to empirical data from the Swiss HIV Cohort Study (SHCS). We used data collected from 1,392 individuals in Switzerland infected with HIV-1 subtype B between 1994 and 2018. The SHCS provided viral load measurements, *pol* gene sequences, and human genotype data for these individuals. We followed the framework outlined above to estimate the pathogen and non-pathogen effects on spVL for the cohort from these data. Figure S2 shows the calculated (total) spVL values, which vary between approximately 1 and 6 log copies/mL in the cohort. Figure S3 shows that this trait is not strongly phylogenetically structured in the cohort, despite high heritability. Finally, figure S4 shows that the estimated non-pathogen effects on spVL correlate quite strongly with total spVL. We estimated spVL heritability in this cohort to be 45% (95% highest posterior density, HPD, 24 - 67%) and selection strength to be 58 time^−1^ (95% HPD 19 - 95) (Figure S5, Table S1). To put these values into the context of our simulation study, they are shown as black points on Figure 2.

We compared our proposed GWAS framework with a more standard approach by performing two different GWAS on the same SHCS human genotypes. In the “GWAS with standard trait value” we used the total trait value, our calculated spVL values, as the GWAS response variable. In the “GWAS with estimated non-pathogen part of trait” we used our estimates for the non-pathogen effects on spVL. Figure 3A shows that results are qualitatively similar between the two GWAS. Q-Q plots show the distribution of p-values are very similar as well (Figure S6). Figure 3B shows how the strength of association changed for some variants in the MHC and *CCR5* regions. Taking into account phylogenetic information slightly decreased association strength for most variants in the *CCR5* region. Association strength increased for some variants in the MHC, for example, SNP rs9265880 had the greatest increase in significance in the MHC region, from a p-value of 3.5 × 10^−07^ to 7.7 × 10^−09^. However, the top-associated variants in the MHC and *CCR5* regions were consistent regardless of the GWAS response variable used (Table S2). Finally Table 1 shows how our GWAS results compare for the two top-associated SNPs identified by McLaren *et al*. (2015). In summary, there are no clear patterns that point to new regions of association in the human genome with spVL when we take into account the pathogen phylogeny.

**Table 1:**
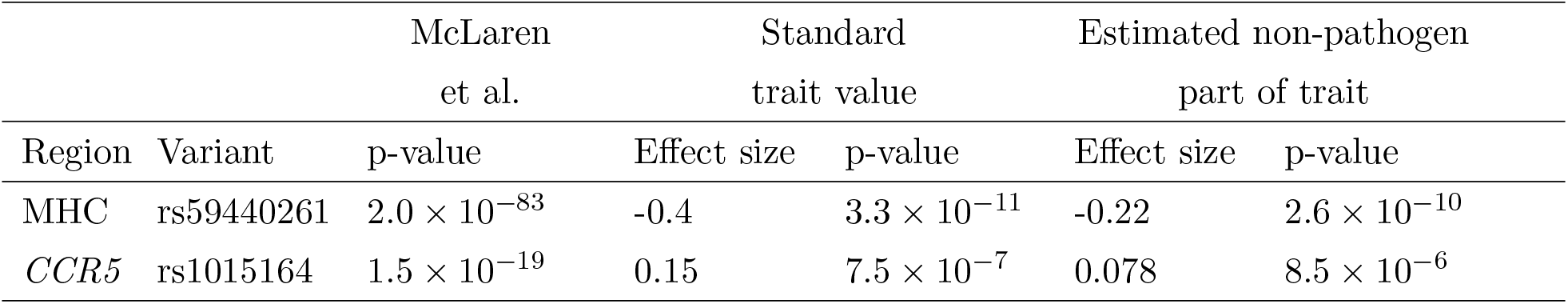
Top association results from McLaren *et al*. (2015) compared to results from this study. Results from this study are for host variants from the SHCS in GWAS with two different response variables. “Standard trait value” means we used the unmodified (total) spVL value and “Estimated non-pathogen part of trait” means we used our estimates for the non-pathogen effects on spVL.

**Figure 3:**
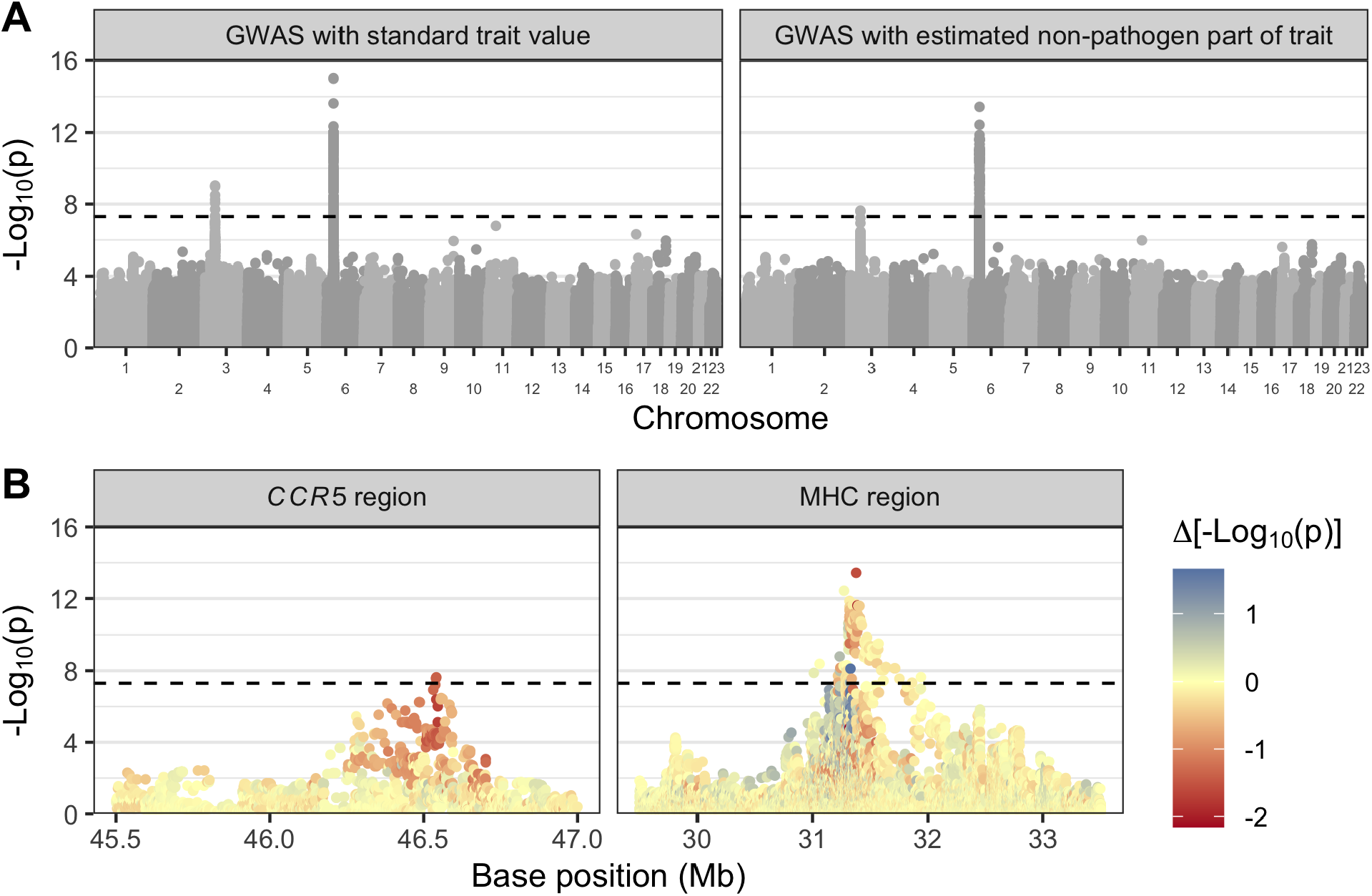
Results from comparative GWAS. (A) shows association p-values for the same host variants from the SHCS cohort in GWAS with two different response variables. On the left, we used unmodified (total) spVL values. On the right, we used our estimates for the non-pathogen effects on spVL. The alternating shades correspond to different chromosomes. (B) compares the strength of association for variants in the *CCR5* and MHC regions between the two GWAS (positions 45.4 - 47Mb on chromosome 3 and 29.5 - 33.5Mb on chromosome 6 for the *CCR5* and MHC, respectively). Base positions are with reference to genome build GRCh37. The color of each point represents the difference in -log_10_ p-value between the two GWAS. Red means taking into account phylogenetic information decreased the strength of association and blue means it increased it. The dashed lines show genome-wide significance at p = 5 × 10^−8^.

## Discussion

In this paper, we presented a new phylogeny-aware GWAS framework to correct for heritable pathogen effects on infectious disease traits. By using information from the pathogen phylogeny, we show it is possible to improve GWAS power to detect host genetic variants associated with a disease trait. This should help us better understand which host factors are protective against a disease versus which increase susceptibility or disease severity.

Our method relies on the POUMM, a model of continuous trait evolution that accounts for heritable and non-heritable effects on a trait, as well as selection. Using this model, we estimated HIV-1 spVL heritability to be 45% (95% HPD 24 - 67%) in the Swiss HIV Cohort Study. Compared to previous studies, this estimate is at the higher end (see Mitov and Stadler (2018) and references therein). Also using the POUMM, Bertels *et al*. (2018) estimated a spVL heritability of 29% (N = 2014, CI 12 - 46%) from the same cohort and Blanquart *et al*. (2017) estimated 31% (N = 2028, CI 15 - 43%) from a pan-European cohort. We note that our sample size (N = 1493 individuals) is smaller than in these other studies. This might be because we restricted samples based on having pol gene sequences with at least 750 non-ambiguous bases. Our aim was to reconstruct a high-quality phylogeny, since the POUMM does not account for phylogenetic uncertainty and the POUMM parameter estimates are key to our downstream trait-correction method. Although our heritability estimate is rather high, the confidence interval largely overlaps that of other studies and we note that estimating heritability per se was not our primary focus.

Instead, the main novelty of our approach was to correct the spVL trait prior to performing the GWAS, thereby estimating and removing pathogen effects. In simulations, we show that when trait heritability amongst infection partners is greater than approximately 25%, GWAS power to detect host genetic variants associated with the same trait is reduced. Our method can correct for this effect in certain evolutionary scenarios by using information from the full pathogen phylogeny.

Based on our simulation results, our method is anticipated to be very useful for disease traits that are highly heritable from donor to recipient and maintain a high correlation between sampled individuals. In simulations, we showed this is the case when heritability is high, selection strength is low, and trait values are not subject to strong stochastic fluctuations. So, cohort-level, phylogenetically structured differences in the measured trait value are necessary for our approach to outperform state of the art methods.

Given our estimates for the heritability of spVL and the selection strength on this trait using Swiss HIV cohort data, our simulation results reveal that we cannot expect a significant improvement in GWAS power for human genetic determinants of spVL (Figure 2). Our method slightly decreases p-values for variants in *CCR5* and slightly decreases some and increases other p-values for variants in the MHC (Figure 3B). Simulations show we shouldn’t expect a net p-value decrease, but our simulations represent an ideal scenario since we simulate under the POUMM. In real life, un-modeled evolutionary pressures like drug treatment and host-specific HLA alleles might cause the reduced p-values. However, the overall picture is consistent between the two GWAS (Figure 3A). Therefore, we conclude that GWAS for host determinants of HIV-1 subtype B spVL is robust to our correction for pathogen effects.

Our method is convenient for GWAS because it is simply a pre-processing step that produces an alternate response variable for GWAS association tests. It is still possible to use previously developed, well-documented, and fast tools for the actual association testing (we used PLINK (Chang *et al*., 2015)). The method relies on the freely available R package POUMM (Mitov and Stadler, 2017) and all the code we wrote is available on the project GitHub at https://github.com/cevopublic/POUMM-GWAS. Future applications of our method might investigate other clinically significant disease traits and outcomes that are affected by both host and pathogen genetic factors, for instance Hepatitis B Virus-related hepatocellular carcinoma (An *et al*., 2018), Hepatitis C treatment success (Ansari *et al*., 2017), and susceptibility to or severity of certain bacterial infections, e.g. Donnenberg *et al*. (2015); Messina *et al*. (2016).

In summary, we argue that infectious disease GWAS should take the pathogen phylogeny into account when searching for host determinants of a disease trait. We give a practical threshold for identifying when GWAS suffer from pathogen effects (heritability of the trait amongst infection partners > 25%) and provide a method that can help in scenarios where trait values are highly heritable and phylogenetically-structured amongst members of a cohort.

## Materials and Methods

### Simulation model

Whenever possible, we tried to parameterize our simulation model for spVL using empirical data. We set the total variance in spVL to 0.73 log copies^2^ mL^−2^ based on UK cohort data (Mitov and Stadler, 2018). Other studies have estimated slightly lower values though (Table S3). After allotting 25% of this variance to the host part of spVL *g*_*h*_ based on results by McLaren *et al*. (2015), we partitioned the remaining variance between the viral part *g*_*v*_ and the environmental partε in different ratios to assess estimator performance across a range of spVL heritabilities. *g*_*h*_ was simulated as the sum of contributions from 20 causal host genetic variants, 10 of which had an effect size of 0.2 log copies mL^−1^ and 10 of which had an effect size of -0.2 log copies mL^−1^. Host genetic variants were generated from a binomial distribution with probability p calculated such that *g*_*h*_ had the appropriate variance (see Table 2). We generated a random viral phylogeny with branch lengths on the same time scale as a previously inferred UK cohort HIV tree (Hodcroft *et al*., 2014) using the R package ape (Paradis and Schliep, 2018). *g*_*v*_ was simulated by running an OU process along the phylogeny using the R package POUMM (Mitov and Stadler, 2017) and sampling values at the tips. For the OU parameters θ and *g*_0_ we used 4.5 log copies mL^−1^ based on fitting the same model to SCHS data (Table S1). This is similar to values previously inferred for HIV (Table S4).

**Table 2:**
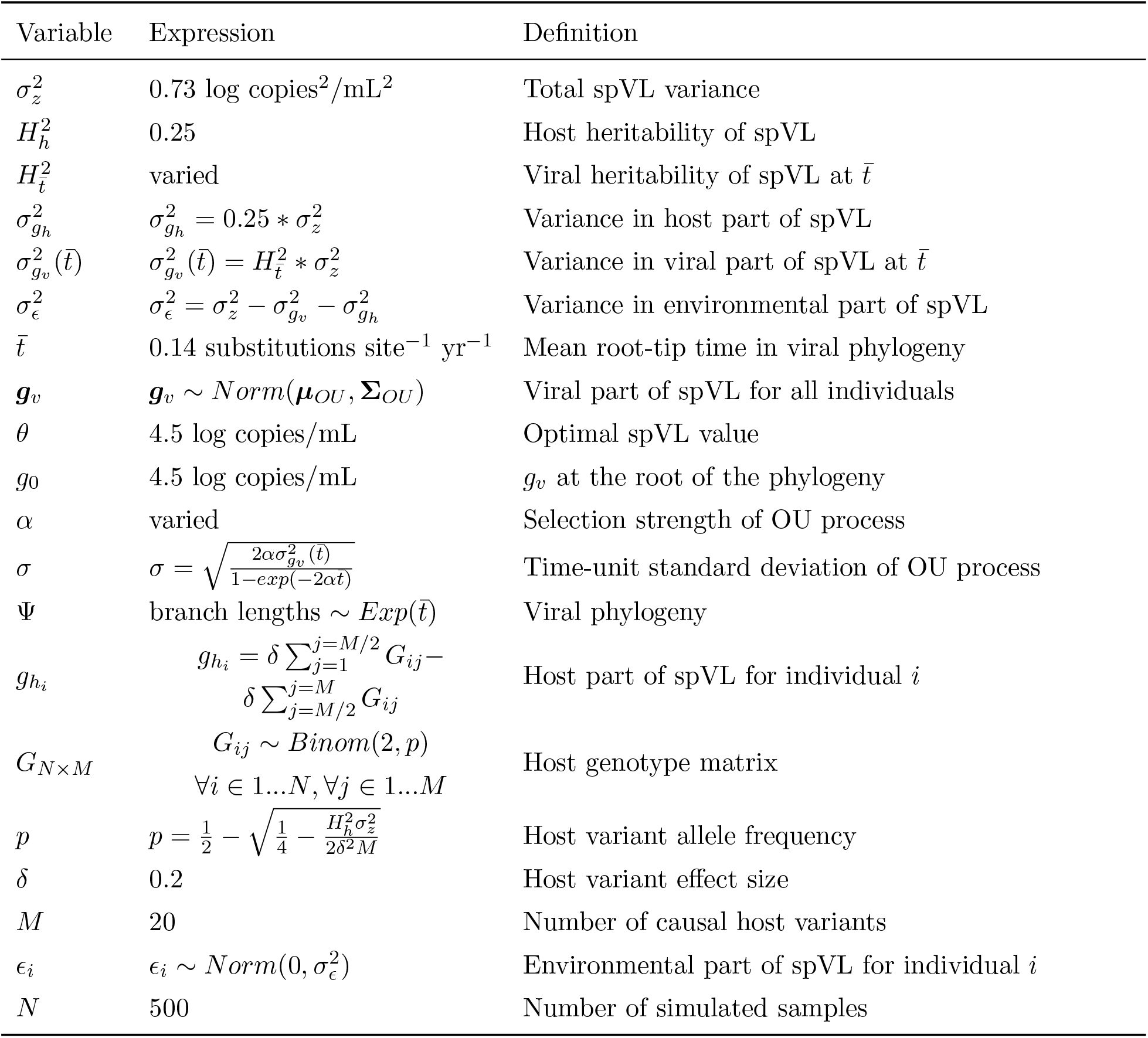
Simulation model parameters. For a full graphical model representation of the simulation scheme, including how these parameters are related, see Figure S1.

To assess our estimator’s performance under a range of evolutionary scenarios, we co-varied the OU parameters for selection strength, *α*, and intensity of random fluctuations, *σ*, so that different proportions of the variability in *g*_*v*_ were attributable to selection and drift, respectively. Finally, the environmental component of spVL *ϵ* was generated from a normal distribution with mean 0. For a full graphical model representation of the simulation scheme, see Figure S1.

### Swiss HIV-1 data

Human genotypes, viral load measurements, and HIV-1 pol gene sequences from HIV-1 positive individuals were all collected in the context of other studies by the Swiss HIV Cohort Study (SHCS) (www.shcs.ch, Scherrer *et al*. (2021); Schoeni-Affolter *et al*. (2010)). All participants were HIV1–infected individuals 16 years or older and written informed consent was obtained from all cohort participants. The anonymized data were made available for this study after the study proposal was approved by the SHCS.

For phylogenetic inference, we retained sequences from 1,493 individuals with non-recombinant subtype B pol gene sequences of at least 750 characters and paired RNA measurements allowing for calculation of spVL, as well as 5 randomly chosen subtype A sequences as an outgroup. We used MUSCLE version 3.8.31 (Edgar, 2004) to align the pol sequences with –maxiters 3 and otherwise default settings. We trimmed the alignment to 1505 characters to standardize sequence lengths. We used IQ-TREE version 1.6.9 (Nguyen *et al*., 2014) to construct an approximate maximum likelihood tree with -m GTR+F+R4 for a general time reversible substitution model with empirical base frequencies and four free substitution rate categories. Otherwise, we used the default IQ-TREE settings. After rooting the tree based on the subtype A samples, we removed the outgroup. Viral subtype was determined by the SHCS using the REGA HIV subtyping tool version 2.0 (de Oliveira *et al*., 2005). We calculated spVL as the arithmetic mean of viral RNA measurements made prior to the start of antiretroviral treatment. For a comparison of several different filtering methods, see Figure S2.

For GWAS, we retained data from 1,392 of the 1,493 SHCS individuals with European ancestry who were not closely related to other individuals in the cohort (Table S5). These were 227 females and 1165 males. Ancestry was determined by plotting individuals along the three primary axes of genotypic variation from a combined dataset of SHCS samples and HapMap populations (Figure S7). Kinship was evaluated using PLINK version 2.3 (Chang *et al*., 2015); we used the –king-cutoff option to exclude one from each pair of individuals with a kinship coefficient > 0.09375. Initial host genotyping quality control and imputation were done as in Thorball *et al*. (2021). Subsequent genotyping quality control was performed using PLINK version 2.3 (Chang *et al*., 2015). We used the options –maf 0.01, –geno 0.01, and –hwe 0.00005 to remove variants with minor allele frequency less than 0.01, missing call rate greater than 0.05, or Hardy-Weinberg equilibrium exact test p-value less than 5×10^−^5. After quality filtering, approximately 6.2 million genetic variants from the 1,392 individuals were retained for GWAS (Table S6).

### POUMM parameter inference

We used the R package POUMM version 2.1.6 (Mitov and Stadler, 2017) to infer the POUMM parameters *g*_0_, *α, θ, σ*, and *σ*_*e*_ from the approximate maximum-likelihood phylogeny and calculated spVL values. The Bayesian inference method implemented in this package requires specification of a prior distribution for each parameter. We used the same, broad prior distributions as in Mitov and Stadler (2018), namely: 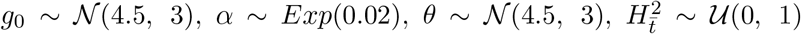, and 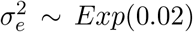. We ran two MCMC chains for 4×10^6^ samples each with a target sample acceptance rate of 0.01 and a thinning interval of 1000. The first 2×10^5^ samples of each chain were used for automatic adjustment of the MCMC proposal distribution. Figure S5 shows the posterior distributions for inferred parameters. Table S1 gives the posterior mean values used for subsequent calculations.

### Phylogenetic spVL correction

We corrected calcualted spVL values using the method described in this paper. For each of the 1,392 individuals in the GWAS cohort, we estimated the viral part of spVL using equation 9 and the corresponding non-viral part using equation 12. For the POUMM parameters *α, σ,θ*, and *σ*_*e*_, we used the posterior mean estimates generated as described above.

### Association testing

We performed two GWAS using the same human genotype data from the SHCS. For the first “GWAS with standard trait value” we used total calculated spVL (*z*) as the response variable for association testing, replicating prior GWAS for host genetic determinants of spVL. For the second “GWAS with estimated non-pathogen part of trait” we replaced total spVL with the estimated nonviral component of spVL (*ê*) as the response variable. Association testing was performed using a linear association model in PLINK version 2.3 (Chang *et al*., 2015) with sex and the top 5 principle components of host genetic variation included as covariates. The sex and principle components covariates were included to reduce residual variance in spVL and control for confounding from host population structure, respectively.

## Data Availability

The simulated data underlying this article can be re-generated using the code available on the project GitHub at https://github.com/cevo-public/POUMM-GWAS. The HIV pathogen genome sequences, clinical data, and human genotypes may be shared on reasonable request to the Swiss HIV Cohort Study at http://www.shcs.ch.

https://github.com/cevo-public/POUMM-GWAS

## Data availability

The simulated data underlying this article can be re-generated using the code available on the project GitHub at https://github.com/cevo-public/POUMM-GWAS. The HIV pathogen genome sequences, clinical data, and human genotypes cannot be shared publicly due to the privacy of individuals who participated in the cohort study. The data may be shared on reasonable request to the Swiss HIV Cohort Study at http://www.shcs.ch.

## Acknowledgments

This work was supported by ETH Zurich. We thank the patients who participate in the SHCS; the physicians and study nurses for excellent patient care; A. Scherrer, E. Mauro, and K. Kusejko from the SHCS Data Centre for data management; and D. Perraudin and M. Amstad for administrative assistance. We also thank Michael Landis, who shared a LaTeX template for graphical model drawing.

The members of the SHCS are: Abela I, Aebi-Popp K, Anagnostopoulos A, Battegay M, Bernasconi E, Braun DL, Bucher HC, Calmy A, Cavassini M, Ciuffi A, Dollenmaier G, Egger M, Elzi L, Fehr J, Fellay J, Furrer H, Fux CA, Günthard HF (President of the SHCS), Hachfeld A, Haerry D (deputy of “Positive Council”), Hasse B, Hirsch HH, Hoffmann M, Hösli I, Huber M, Kahlert CR (Chairman of the Mother Child Substudy), Kaiser L, Keiser O, Klimkait T, Kouyos RD, Kovari H, Kusejko K (Head of Data Centre), Martinetti G, Martinez de Tejada B, Marzolini C, Metzner KJ, Müller N, Nemeth J, Nicca D, Paioni P, Pantaleo G, Perreau M, Rauch A (Chairman of the Scientific Board), Schmid P, Speck R, Stöckle M (Chairman of the Clinical and Laboratory Committee), Tarr P, Trkola A, Wandeler G, Yerly S.

The Swiss HIV Cohort Study is supported by the Swiss National Science Foundation (grant 201369), by SHCS project 858 and by the SHCS research foundation. Furthermore, the SHCS drug resistance database is supported by the Yvonne Jacob Foundation (to HFG). The data are gathered by the Five Swiss University Hospitals, two Cantonal Hospitals, 15 affiliated hospitals and 36 private physicians (listed in http://www.shcs.ch/180-health-care-providers).

