## Supplementary materials for "A phylogeny-aware GWAS framework to correct for heritable pathogen effects on infectious disease traits"

### Supplemental Material

#### Expected results from simulations

Here we show the root mean square error (RMSE) of $\boldsymbol{z} - \bar{\boldsymbol{z}}$ as an estimate for $\boldsymbol{h}$ should be $\approx 0.74$ in our simulation scheme. First we write the expression for the RMSE:

$$RMSE = \sqrt{\frac{\sum_i^N (z - \bar{z} - h)^2}{N}} \tag{13}$$

Note that $z - \bar{z}$ differs from $h$ due to a viral effect and an environmental effect. So the term inside the square root equals the combined variance of these two effects:

$$RMSE = \sqrt{\sigma_{g_v}^2 + \sigma_\epsilon^2} \tag{14}$$

We can calculate the variance due to these two effects because the total variance in spVL $\sigma_z^2$, and the fraction of the total variance due to host genetic effects, $\sigma_{g_h}^2$, are fixed parameters in our simulation scheme.

$$\begin{aligned} \sigma_{g_h}^2 + \sigma_{g_v}^2 + \sigma_\epsilon^2 &= \sigma_z^2 \\ 0.25 * \sigma_z^2 + \sigma_{g_v}^2 + \sigma_\epsilon^2 &= \sigma_z^2 \\ \sigma_{g_v}^2 + \sigma_\epsilon^2 &= 0.75 * \sigma_z^2 \\ \sigma_{g_v}^2 + \sigma_\epsilon^2 &= 0.75 * 0.73 \\ \sigma_{g_v}^2 + \sigma_\epsilon^2 &= 0.55 \end{aligned} \tag{15}$$

Therefore, we can expect the RMSE for $\boldsymbol{z} - \bar{\boldsymbol{z}}$ as an estimate for $\boldsymbol{h}$ to be around $\sqrt{0.55} \approx 0.74$.

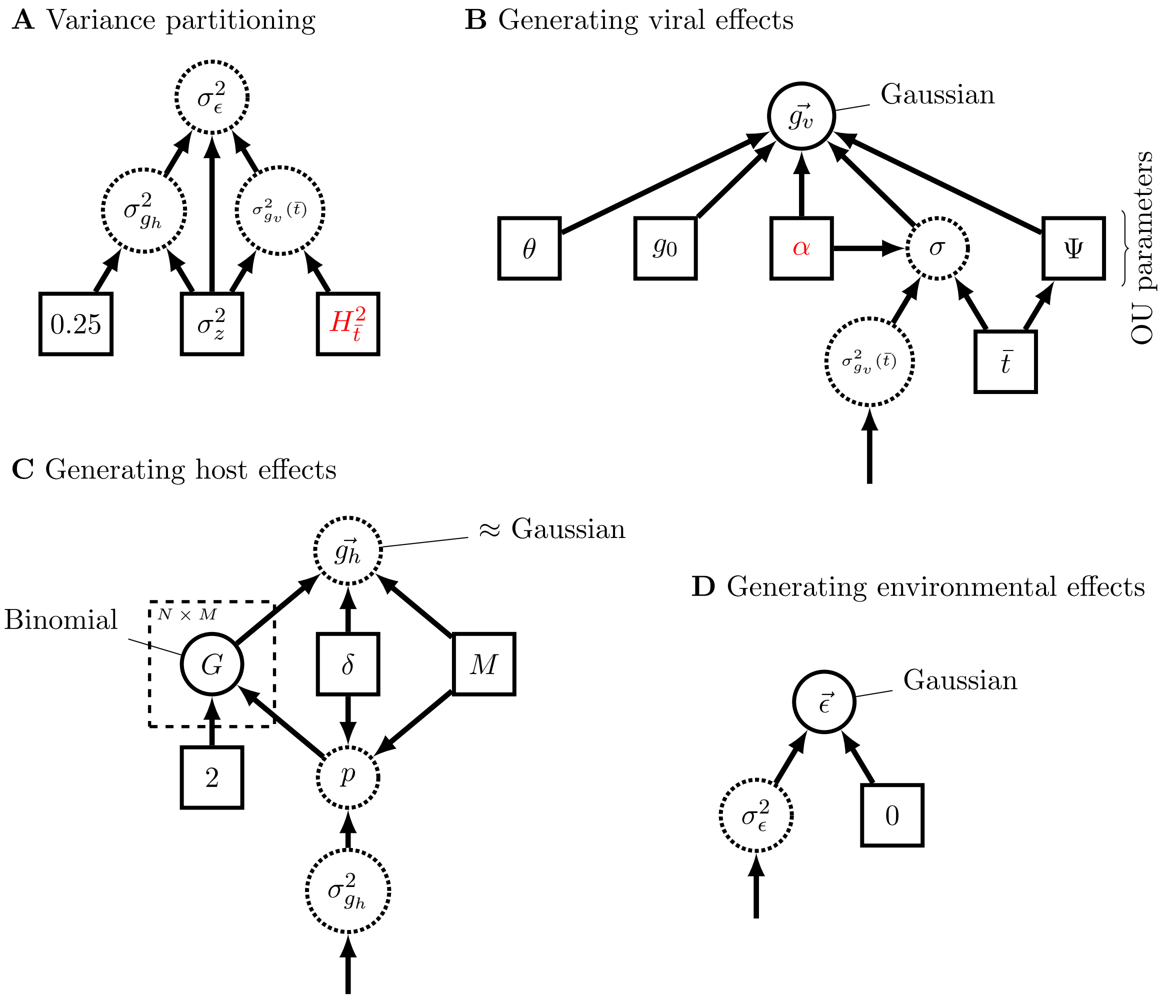


Figure S1: A graphical model representation of our simulation scheme, following the recommendations in Höhna *et al.* (2014). Variables in solid squares are constants, with the two master control variables that we vary from simulation to simulation highlighted in red. Variables in solid circles are realizations of random variables and variables in dashed circles are determined as a function of other variables. Arrows represent dependencies among variables and the dashed square represents repetition. All parameters are defined in Table 2, as well as the values or expressions used for them. (A) shows how the variance in the simulated environmental effect $\sigma^2_\epsilon$ is smaller if the master pathogen heritability value $H^2_{\bar{t}}$ is higher and vice-versa. (B) shows the OU parameters and the pathogen phylogeny, which generate the Gaussian-distributed pathogen effects. The OU parameters $\theta$ and $g_0$ are fixed, whereas $\sigma$ is a deterministic function of the variance in the pathogen effect and the value of $\alpha$. In other words, we use $\sigma$ to maintain the desired pathogen heritability while varying $\alpha$. We generate a new random phylogeny for each simulation. (C) shows how host genotypes are drawn to generate host effects. The host genotype matrix $G$ contains the number of copies (0, 1, or 2) for each of $M$ causal variants with effect size $\delta$. We assume half the variants have a positive effect and half have a negative effect. The allele frequency $p$ for the causal variants set so that we achieve the desired variance in the host effects. (D) shows that the environmental effect is drawn from a Gaussian distribution with mean zero and variance as determined in part (A).

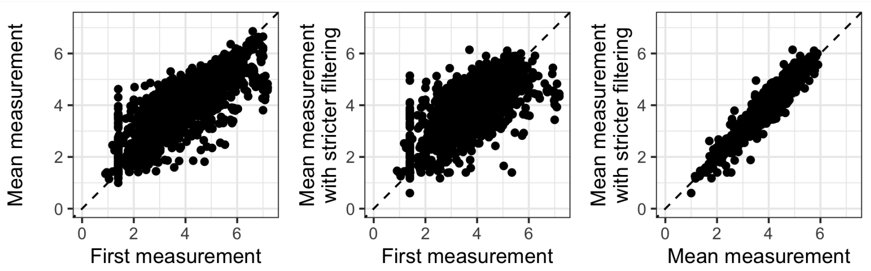


Figure S2: A comparison of different ways to calculate spVL based on viral load measurements provided by the SHCS. The stricter filtering excludes all measurements possibly $< 6$ months after infection and after treatment or AIDS, whereas the more lenient filtering excludes only measurements after treatment. We used the lenient filter, mean measurement values because these correlate well with the values from the stricter filter but allow us to retain many more individuals from the cohort for our study.

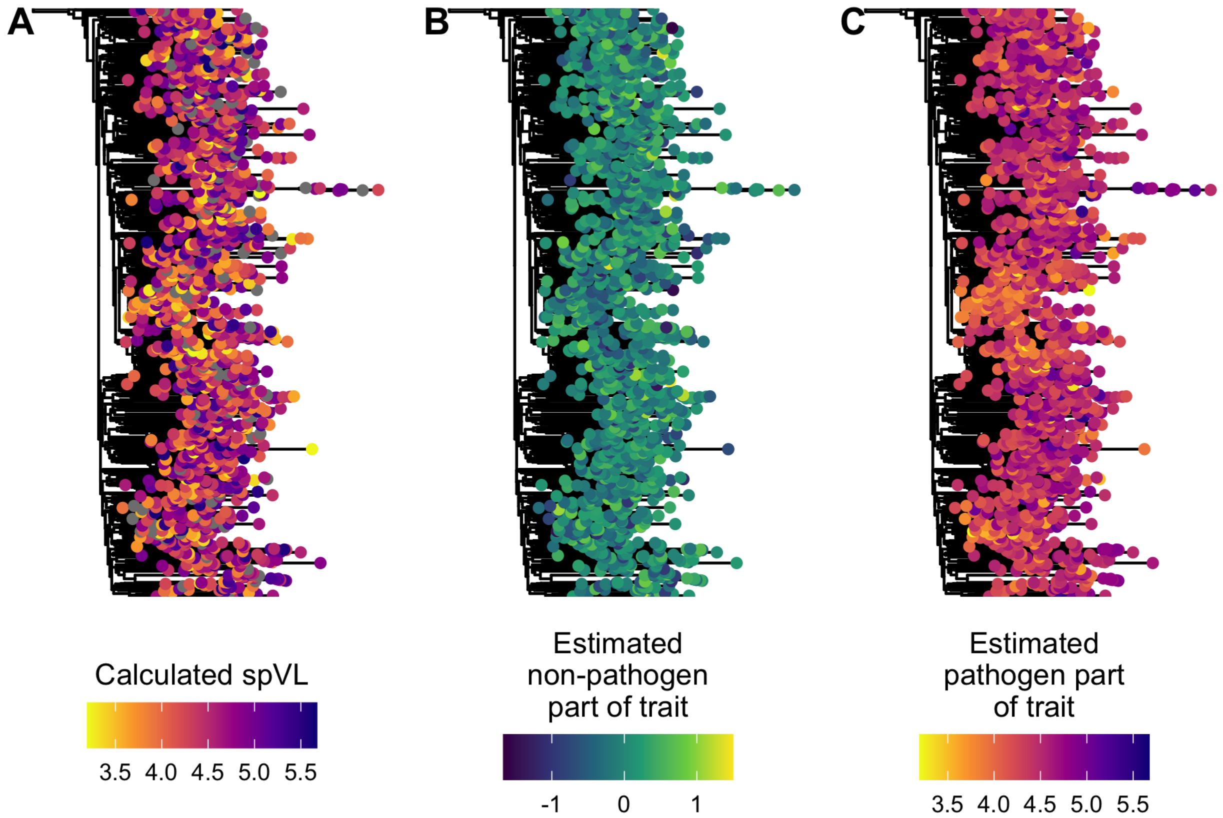


Figure S3: Inferred HIV-1 *pol* gene phylogeny with tips colored by (A) calculated spVL, (B) estimated non-pathogen effects on spVL and (C) estimated pathogen effects on spVL.

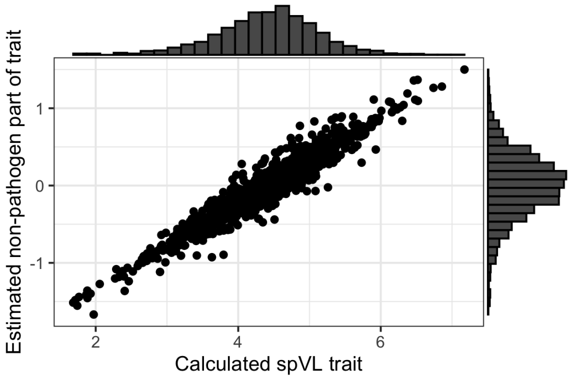


Figure S4: A comparison of measured (calculated) spVL values versus our estimated non-pathogen effect on spVL for each SHCS cohort member used in the study. The histograms show the marginal distribution of each value across the individuals.

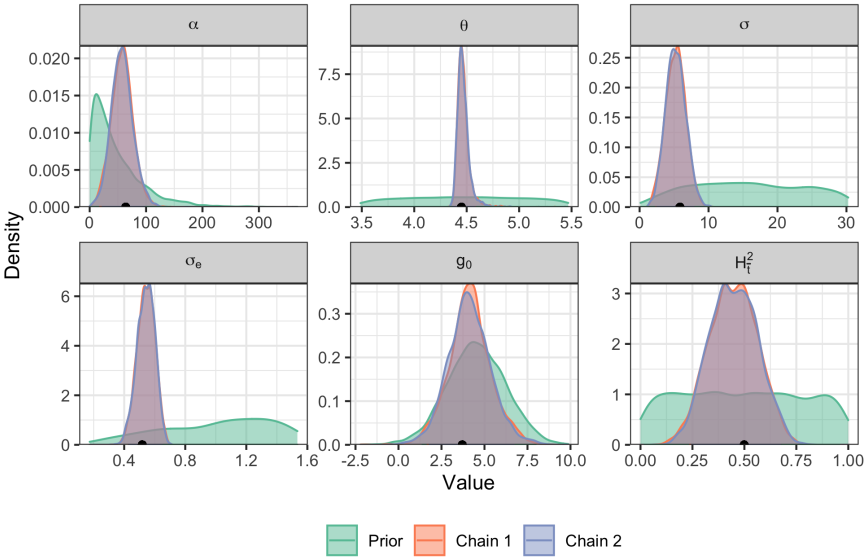


Figure S5: Posterior distributions compared to the prior for POUMM parameter estimates based on SHCS data. We ran two different MCMC chains to ensure the estimates converged. The black point on the x-axis shows the posterior mean value, which was used to estimate the pathogen- and non-pathogen effects on spVL.

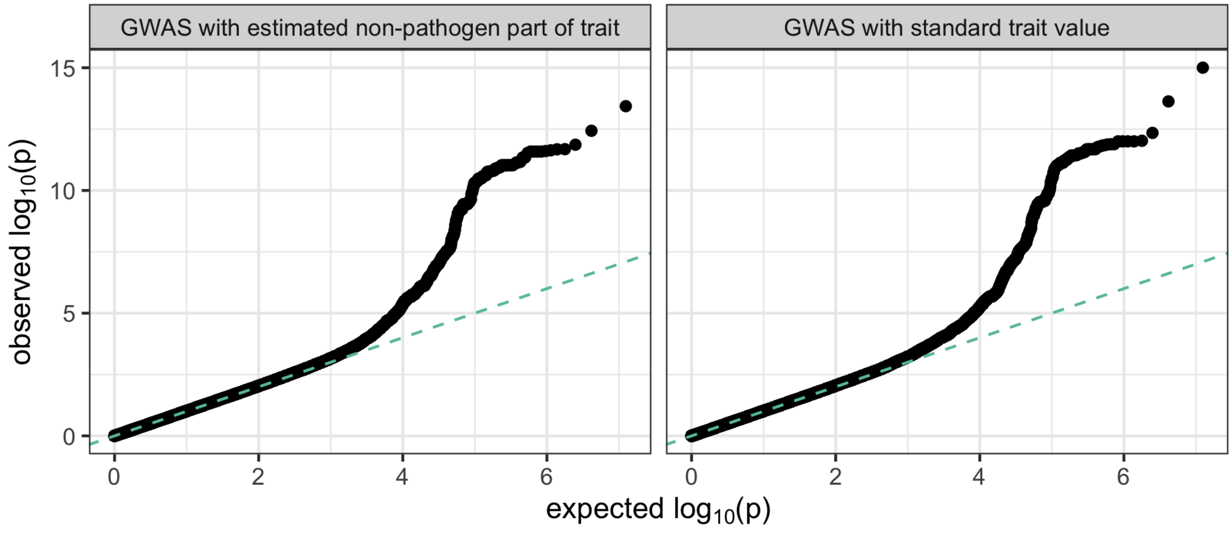


Figure S6: Quartile-quartile plots from association tests. The dashed green line shows the y = x line.

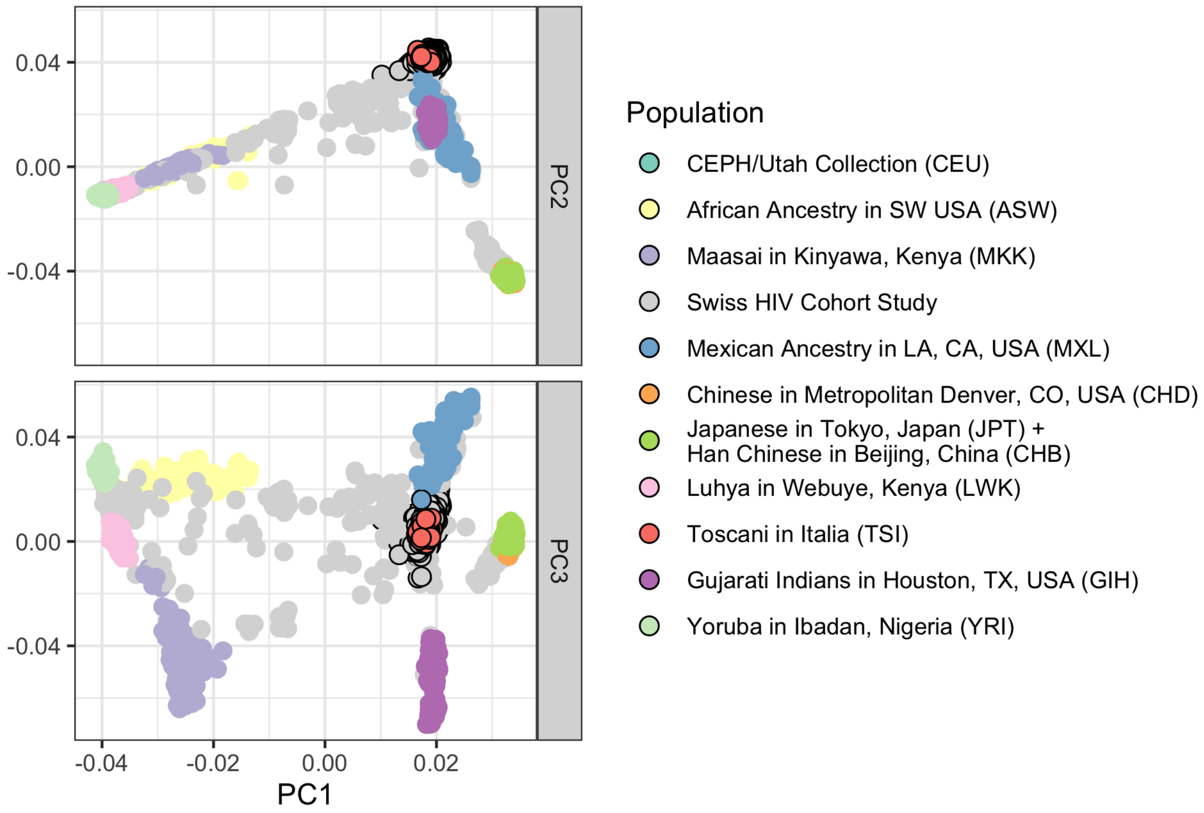


Figure S7: SHCS individuals and HapMap3 individuals plotted along the top three principle components of genetic variation. Points with black borders are within the thresholds used to select individuals of likely European ancestry.

Table S1: POUMM parameter estimates for spVL based on SHCS data. HPD = Highest posterior density.

| Parameter | Posterior mean | 95% HPD |
|---|---|---|
| $g_0$ | 4.23 | (1.72, 6.71) |
| $\theta$ | 4.47 | (4.37, 4.58) |
| $\sigma$ | 5.25 | (2.37, 7.9) |
| $\alpha$ | 57.65 | (19.49, 95.2) |
| $\sigma_e$ | 0.54 | (0.43, 0.65) |
| $H_t^2$ | 0.45 | (0.24, 0.67) |

Table S2: Effect size and p-values from the top most strongly associated variants in the *CCR5* and MCR regions from each of the two GWAS performed in our study. "Standard" means the GWAS with standard spVL trait values and "Corrected" means the GWAS with the estimated non-pathogen part of the trait. Entries above the dividing line are the top-associated variants from the "Standard" GWAS and entries below the dividing line are the top-associated variants from the "Corrected" GWAS. Many entries overlap between the two.

| Region | Position | Variant | Standard effect size | Standard p-value | Corrected effect size | Corrected p-value |
|---|---|---|---|---|---|---|
| *CCR5* | 46531144 | rs9845968 | -0.16 | $5.6 \times 10^{-9}$ | -0.083 | $1.2 \times 10^{-7}$ |
| *CCR5* | 46537849 | rs867620 | -0.16 | $3.2 \times 10^{-9}$ | -0.085 | $6 \times 10^{-8}$ |
| *CCR5* | 46539864 | rs11130092 | -0.16 | $1.1 \times 10^{-9}$ | -0.087 | $2.6 \times 10^{-8}$ |
| *CCR5* | 46540932 | rs10865942 | -0.16 | $8.4 \times 10^{-9}$ | -0.081 | $4 \times 10^{-7}$ |
| *CCR5* | 46541147 | rs7430431 | -0.17 | $9.2 \times 10^{-10}$ | -0.088 | $2.3 \times 10^{-8}$ |
| MHC | 31274380 | rs9264942 | -0.21 | $4.5 \times 10^{-13}$ | -0.12 | $3.7 \times 10^{-13}$ |
| MHC | 31321919 | rs1055821 | -0.33 | $9.4 \times 10^{-13}$ | -0.19 | $1.4 \times 10^{-12}$ |
| MHC | 31380034 | rs112243036 | -0.32 | $9.9 \times 10^{-16}$ | -0.17 | $3.7 \times 10^{-14}$ |
| MHC | 31391401 | rs4418214 | -0.34 | $2.4 \times 10^{-14}$ | -0.18 | $2.5 \times 10^{-12}$ |
| MHC | 31400137 | rs138130755 | -0.46 | $1 \times 10^{-12}$ | -0.26 | $2.6 \times 10^{-12}$ |
| MHC | 31400705 | rs138117378 | -0.46 | $1 \times 10^{-12}$ | -0.26 | $2.6 \times 10^{-12}$ |
| MHC | 31402358 | rs148792134 | -0.46 | $1 \times 10^{-12}$ | -0.26 | $2.6 \times 10^{-12}$ |
| MHC | 31409677 | rs140991764 | -0.46 | $1 \times 10^{-12}$ | -0.26 | $2.6 \times 10^{-12}$ |
| *CCR5* | 46531144 | rs9845968 | -0.16 | $5.6 \times 10^{-9}$ | -0.083 | $1.2 \times 10^{-7}$ |
| *CCR5* | 46537849 | rs867620 | -0.16 | $3.2 \times 10^{-9}$ | -0.085 | $6 \times 10^{-8}$ |
| *CCR5* | 46539864 | rs11130092 | -0.16 | $1.1 \times 10^{-9}$ | -0.087 | $2.6 \times 10^{-8}$ |
| *CCR5* | 46541147 | rs7430431 | -0.17 | $9.2 \times 10^{-10}$ | -0.088 | $2.3 \times 10^{-8}$ |
| *CCR5* | 46556835 | rs6808142 | 0.15 | $8.3 \times 10^{-8}$ | 0.082 | $3.3 \times 10^{-7}$ |
| MHC | 31274380 | rs9264942 | -0.21 | $4.5 \times 10^{-13}$ | -0.12 | $3.7 \times 10^{-13}$ |
| MHC | 31321919 | rs1055821 | -0.33 | $9.4 \times 10^{-13}$ | -0.19 | $1.4 \times 10^{-12}$ |
| MHC | 31367874 | rs111281598 | -0.37 | $1.5 \times 10^{-12}$ | -0.22 | $2.1 \times 10^{-12}$ |
| MHC | 31376266 | rs73400361 | -0.37 | $1.4 \times 10^{-12}$ | -0.22 | $2.1 \times 10^{-12}$ |
| MHC | 31380034 | rs112243036 | -0.32 | $9.9 \times 10^{-16}$ | -0.17 | $3.7 \times 10^{-14}$ |

Table S3: Summary statistics for log spVL in previously sampled populations. $\bar{z}$ is average spVL (log copies/mL) and $\sigma_z^2$ is variance in measured spVL ( log copies$^2$/mL$^2$). Values from (Blanquart *et al.*, 2017; Mitov and Stadler, 2018) are empirical; values from (Bonhoeffer *et al.*, 2015) were estimated by fitting a normal distribution to the data.

| Measurement | Value | Reference |
|---|---|---|
| $\bar{z}$ | $\approx$ 4.5 | Mitov and Stadler (2018) |
| $\bar{z}$ | 4.4 | Blanquart *et al.* (2017) |
| $\bar{z}$ | $\approx$ 4.5 | Bonhoeffer *et al.* (2015) |
| $\sigma_z^2$ | 0.73 | Mitov and Stadler (2018) |
| $\sigma_z^2$ | 0.50 | Blanquart *et al.* (2017) |
| $\sigma_z^2$ | $\approx$ 0.5 | Bonhoeffer *et al.* (2015) |

Table S4: POUMM parameter estimates for spVL from previous studies.

| Parameter | Value (Uncertainty) | Reference | Notes |
|---|---|---|---|
| $g_0$ | 5.54 (4.04 - 7.25) | Mitov and Stadler (2018) | 8,483 UK HIV cohort individuals, *pol* tree |
| $\theta$ | 4.45 (4.41 - 4.49) | Mitov and Stadler (2018) | |
| $\theta$ | 4.0 (1.6 - 4.)) | Bertels *et al.* (2018) | 3,036 SHCS individuals, *pol* tree |
| $\theta$ | 4.1 (3.5 - 4.9) | Blanquart *et al.* (2017) | 1,581 subtype B individuals from Europe, whole genome tree |
| $\alpha$ | 28.78 (16.64 - 46.93) | Mitov and Stadler (2018) | |
| $\alpha$ | 32.7 (0.03 - 57.6) | Bertels *et al.* (2018) | |
| $\alpha$ | 7.6 (1.2 - 10) | Blanquart *et al.* (2017) | **limited $\alpha$ to $\leq$ 10 |
| $\sigma$ | 2.97 (1.95 - 4.37) | Mitov and Stadler (2018) | |
| $\sigma$ | 1.3 (0.66 -1.87) | Blanquart *et al.* (2017) | |
| $\sigma_e$ | 0.77 (0.73, 0.8) | Mitov and Stadler (2018) | |
| $\sigma_e$ | 0.61 (0.54, 0.65) | Blanquart *et al.* (2017) | |

Table S5: Number of samples for GWAS after sequential filtering steps.

| Sample filter | Number of samples remaining |
|---|---|
| Subtype B pol sequences | 1516 |
| With paired spVL measurement | 1516 |
| $>$ 750 characters in sequence | 1493 |
| Individual is of European ancestry | 1396 |
| Kinship coefficient $> 0.09375$ | 1392 |

Table S6: Number of variants for GWAS after sequential filtering steps.

| Variant filter | Number of variants remaining |
|---|---|
| Raw data | 76979521 |
| Missing genotype rate $> 0.05$ | 11590002 |
| Hardy-Weinburg exact test p-value $< 5 \times 10^{-5}$ | 11589246 |
| Minor allele frequency $< 0.01$ | 6228626 |
